# Short-Term Relative Effectiveness of Homologous NVX-CoV2373 and BNT162b2 COVID-19 Vaccinations in South Korea

**DOI:** 10.1101/2024.07.02.24309830

**Authors:** Eunseon Gwak, Seung-Ah Choe, Erdenetuya Bolormaa, Young June Choe, Chengbin Wang, Jonathan Fix, Muruga Vadivale, Matthew D. Rousculp

## Abstract

To estimate the relative, short-term effectiveness of NVX-CoV2373 versus BNT162b2 (Pfizer– BioNTech) in preventing SARS-CoV-2 infection and severe COVID-19 disease during the Omicron variant dominance in South Korea. Retrospective cohort-study among ≥12-year-olds using the K-COV-N database, which links COVID-19 vaccine registry data with health insurance claims data. The Cox proportional-hazards model and inverse probability of treatment weighting were employed to calculate adjusted hazard ratios (aHRs). Among homologous primary-series NVX-CoV2373 versus BNT162b2 recipients, the aHR was 0.86 (95% CI: 0.82–0.91) for all laboratory-confirmed and 0.80 (95% CI: 0.48–1.33) for severe infections. Among homologous 1st-booster recipients, it was 0.74 (95% CI: 0.61–0.90) for all laboratory-confirmed and 0.48 (95% CI: 0.12–1.92) for severe infections. At 30-days post-immunization, we observed homologous, NVX-CoV2373 primary-series and 1st-booster offered added protection against SARS-CoV-2 infection versus BNT162b2; while there was numerically lower risk of severe disease among NVX-CoV2373-vaccinated, no statistically significant differences were observed.

## Introduction

NVX-CoV2373, a monovalent protein-based SARS-CoV-2 vaccine demonstrated high efficacy against coronavirus disease (COVID-19) in the United States and Mexico (1). As NVX-CoV2373 is globally integrated into national immunization programs, including South Korea, it is imperative to assess its real-world effectiveness against COVID-19. While evidence indicates NVX-CoV2373 induces similar neutralizing antibody titers as COVID-19 mRNA vaccines, there is limited data on its relative effectiveness in the general population (2). This study aims to estimate the relative short-term effectiveness of NVX-CoV2373 compared to BNT162b2 (Pfizer–BioNTech) in preventing SARS-CoV-2 infection and critical COVID-19 during the Omicron variant dominance in South Korea, using population-based data.

## Methods

We used the Korea Disease Control and Prevention Agency-COVID-19-National Health Insurance Service (K-COV-N) cohort database that links the COVID-19 vaccine registry data with the health insurance claims data (3). Vaccination status includes vaccination date, vaccine type and dose, and place of vaccination. We then constructed a cohort to compare the risk of all laboratory-confirmed and severe SARS-CoV-2 infections (intensive care unit [ICU] admission due to COVID-19 or death within 30-days of SARS-CoV-2 infection) among the NVX-CoV2373 vaccine recipients compared with BNT162b2 recipients between February and December 2022. Because the cause of death was unavailable in the data, death after SARS-CoV-2 infection within the risk window was assumed to be related to it. The inclusion criteria capture all persons ≥12 years with known COVID-19 vaccination status, complete medical outcome data, at least 365-days of follow-up before the index date (date of receiving a dose of interest), and residing in South Korea. Our analysis was restricted only to individuals completing homologous primary or booster series immunization.

We estimated propensity scores (PS) for receiving NVX-CoV2373 based on relative income level (medical aid beneficiary and income quartiles), residential region (Seoul capital area or not), disability registration, prior COVID-19 infection history, Charlson comorbidity index assessed in the previous year, and month of the latest immunization. We used PS-based inverse probability of treatment weighting (IPTW) to control for confounding by balancing patient characteristics at baseline. We trimmed the tails of the IPTW distribution observations below a PS of 5% and above a PS of 95% of the vaccinated cohorts, thereby excluding those with extreme values (4). To compare vaccine effectiveness (VE), we estimated the incidence rate (IR per 1000 person-days) and IPTW-adjusted hazard ratios (aHR) of all laboratory-confirmed and severe SARS-CoV-2 infections considering time-varying exposures to vaccination.

The risk window for outcomes was 30-days (NOTE: The risk window started either after 7 days (booster) or 14 days (primary series) post-vaccination). For each outcome, individuals were followed for the duration of the risk window until the earliest of one of the following: documentation of the COVID-19 morbidity, death, maximum follow-up duration (30-days), next dose administration, and the end of the study calendar period (December 31, 2022).

The relative VE of NVX-CoV2373 vaccination versus BNT162b2 was assessed regarding overall and severe SARS-CoV-2 infections. The Cox proportional-hazards model was employed to calculate the aHR of SARS-CoV-2 infection, and severe COVID-19 for NVX-CoV2373 compared with BNT162b2. All statistical analyses were performed using SAS^®^ 9.4 (SAS Institute, Cary, NC, USA). This study was approved by the Korea University Institutional Review Board (IRB No. 2023AN0124).

## Results

Among the 137,845 recipients (104,004 for NVX-CoV2373 and 33,841 BNT162b2) of homologous primary series, 17,134 and 4439 received homologous 1^st^ booster doses, and 1,177 and 363 received homologous 2^nd^ booster doses, respectively (Table 1). The NVX-CoV2373 recipients were older, more likely to be male, have a history of prior SARS-CoV-2 infection, have a disability, be employed, and were found to have more comorbidities (CCI ≥3), and higher income level.

**Table 1.** General characteristics of the population who received homologous series of NVX-CoV2373 and BNT162b2 vaccines in South Korea, from February to December 2022.

| Variables | Primary series |  |  | 1st homologous booster |  |  | 2nd homologous booster |  |  |
| --- | --- | --- | --- | --- | --- | --- | --- | --- | --- |
|  | NVX-CoV2373 | BNT162b2 | *P* | NVX-CoV2373 | BNT162b2 | *P* | NVX-CoV2373 | BNT162b2 | *P* |
| N | 104,004 | 33,841 |  | 17,134 | 4439 |  | 1177 | 363 |  |
| Age, median | 44 (IQR: 26) | 36 (IQR: 23) | <0.001 | 57 (IQR: 29) | 57 (IQR: 37) |  | 68 (IQR: 16) | 71 (IQR: 16) |  |
| Sex |  |  |  |  |  |  |  |  |  |
| Male | 45,102 (43.4) | 8293 (24.5) | <0.001 | 7767 (45.3) | 1373 (30.9) | <0.001 | 631 (53.6) | 211 (58.1) | 0.131 |
| Female | 58,902 (56.6) | 25,548 (75.5) |  | 9367 (54.7) | 3066 (69.1) |  | 546 (46.4) | 152 (41.9) |  |
| Non-capital area <sup>a</sup> | 52,397 (50.4) | 17,163 (50.7) | 0.282 | 8061 (47.1) | 2072 (46.7) | 0.66 | 506 (43) | 167 (46) | 0.311 |
| Prior infection | 80,559 (77.5) | 21,391 (63.2) | <0.001 | 15,990 (93.3) | 4003 (90.2) | <0.001 | 1175 (99.8) | 355 (97.8) | <0.001 |
| Disability | 7896 (7.5) | 2078 (6.1) | <0.001 | 2290 (13.4) | 604 (13.6) | 0.674 | 213 (18.1) | 89 (24.5) | 0.007 |
| CCI ≥3 | 14,503 (13.9) | 3312 (9.8) | <0.001 | 4000 (23.4) | 924 (20.8) | <0.001 | 461 (39.2) | 122 (33.6) | 0.056 |
| Employed | 34,378 (33.1) | 7781 (23.0) | <0.001 | 4362 (25.5) | 683 (15.4) | <0.001 | 185 (15.7) | 25 (6.9) | <0.001 |
| Income level <sup>b</sup> |  |  |  |  |  |  |  |  |  |
| 1Q (lowest) | 4750 (4.7) | 1732 (5.4) | <0.001 | 1385 (8.3) | 537 (12.5) | <0.001 | 138 (11.9) | 79 (21.9) | <0.001 |
| 2Q | 14,456 (14.4) | 4107 (12.9) |  | 2450 (14.6) | 631 (14.7) |  | 155 (13.4) | 59 (16.4) |  |
| 3Q | 20,019 (19.9) | 6761 (21.2) |  | 2988 (17.9) | 834 (19.5) |  | 184 (15.9) | 61 (16.9) |  |
| 4Q | 25,477 (25.3) | 9783 (30.7) |  | 3839 (23.0) | 1111 (26.0) |  | 222 (19.1) | 73 (20.3) |  |
| 5Q (richest) | 35,985 (35.7) | 9455 (29.7) |  | 6069 (36.3) | 1168 (27.3) |  | 462 (39.8) | 88 (24.4) |  |

<sup>a</sup> Non-capital area denotes residence outside of Seoul, Incheon metropolitan city, and Gyeonggi province, included in the Seoul Capital Area of South Korea.
<sup>b</sup> Income levels, 1Q, 1<sup>st</sup> quintile (lowest); 2Q, 2<sup>nd</sup> quintile, 3Q, 3<sup>rd</sup> quintile; 4Q, 4<sup>th</sup> quintile, 5Q, 5<sup>th</sup> quintile (richest); values missing for 5320 individuals in primary series, 561 in 1<sup>st</sup> booster, and 19 in 2<sup>nd</sup> booster vaccination groups.
Abbreviations: CCI, Charlson comorbidity index; IQR, interquartile range

Table 2 showed IR and aHR of all laboratory-confirmed and severe SARS-CoV-2 infections following primary series, 1^st^ booster, and 2^nd^ booster of NVX-CoV2373 or BNT162b2 homologous doses. Among the homologous NVX-CoV2373 primary series recipients, 4884 (IR=1.53, 95% CI: 1.48–1.57) experienced an infection, with 58 (IR=0.02; 95% CI: 0.01–0.02) having a severe infection. Among BNT162b2 primary series recipients, 4,249 (IR=4.36; 95% CI: 4.23–4.49) had laboratory-confirmed infections, and 34 (IR=0.03; 95% CI: 0.02–0.05) experienced a severe infection. Comparing NVX-CoV2373 to BNT162b2, the aHR was 0.86 (95% CI: 0.82–0.91) for all laboratory-confirmed and 0.80 (95% CI: 0.48–1.33) for severe infections. Among homologous 1^st^ booster recipients, the aHR was 0.74 (95% CI: 0.61–0.90) for all laboratory-confirmed and 0.48 (95% CI: 0.12–1.92) for severe infections.

**Table 2.** Incidence rates (per 1000 person-days) and adjusted hazard ratios, after inverse probability of treatment weighting based on propensity scores, of all laboratory-confirmed and severe SARS-CoV-2 infections among NVX-CoV2373 and BNT162b2 vaccinations in South Korea, from February to December 2022.

| Dose | NVX-CoV2373 |  |  | BNT162b2 |  |  | aHR (95% CI)<sup>b</sup> |
| --- | --- | --- | --- | --- | --- | --- | --- |
|  | Event | Person-days | IR (95% CI) | Event | Person-days | IR (95% CI) |  |
| Primary series |  |  |  |  |  |  |  |
| All lab-confirmed infections | 4884 | 3,029053 | 1.53 (1.48–1.57) | 4249 | 957,188 | 4.36 (4.23–4.49) | 0.86 (0.82–0.91) |
| Severe infections<sup>a</sup> | 50 | 3,119,176 | 0.02 (0.01–0.02) | 27 | 1,014,824 | 0.03 (0.02–0.05) | 0.80 (0.48–1.33) |
| 1<sup>st</sup> booster |  |  |  |  |  |  |  |
| All lab-confirmed infections | 497 | 506,049 | 0.94 (0.86–1.03) | 98 | 131,541 | 0.75 (0.57–0.86) | 0.74 (0.61–0.90) |
| Severe infections | 6 | 513,944 | 0.01 (0.01–0.03) | 3 | 133,128 | 0.02 (0.01–0.07) | 0.48 (0.12–1.92) |
| 2<sup>nd</sup> booster |  |  |  |  |  |  |  |
| All lab-confirmed infections | 1 | 35,230 | 0.03 (0.00–0.20) | 6 | 10,810 | 0.56 (0.25–1.24) | 0.62 (0.01-59.02) |
| Severe infections | 0 | 35,310 | 2.11E-13(0,infty) | 1 | 10,884 | 0.09 (0.01–0.65) | Unable to assess |

<sup>a</sup> Severe SARS-CoV-2 infection was defined as intensive care unit (ICU) admission due to COVID-19 and death within 30 days of post-SARS-CoV-2 infection.
<sup>b</sup> HR was calculated using inverse probability of treatment weighting (IPTW) based on the propensity score for receiving NVX-CoV2373, with IPTW adjustment for relative level of income, residential region (Seoul capital area or not), disability registration, prior COVID-19 infection history, and the Charlson comorbidity index assessed before the date of latest immunization.
Abbreviation, aHR, IPTW-adjusted hazard ratio; CI, confidence interval; ICU, intensive care unit; IR, incidence rate.

## Discussion

In this study, we observed that during the Omicron variant predominance in South Korea, NVX-CoV2373 primary series offered added protection against laboratory-confirmed SARS-CoV-2 infection than BNT162b2; while lower risk in NVX-CoV2373 in preventing severe infection, but no statistical significance were observed. Comparable protection was observed after the 1^st^ booster doses of NVX-CoV2373 with BNT162b2, and no comparison was assessed on the 2^nd^ booster dose due to a lack of events after receiving the 2^nd^ booster dose of NVX-CoV2373 (Table 2). Our finding contrasts with a previous efficacy meta-analysis (5). Nonetheless, our data represent the initial instance demonstrating the variation in effectiveness for homologous doses (2 vs. 2, 3 vs. 3, and 4 vs. 4) of NVX-CoV2373 and BNT162b2 in a vast population-based database.

Several limitations should be considered when interpreting the results. This study relied on health insurance data and may lack information on all confounders, and the variant of previous infections. Furthermore, time since previous infection might have been different across NVX-CoV2373 and BNT162b2, However, the medical guidance in South Korea at the time was to delay receipt of COVID-19 vaccine until 3 months after a confirmed SARS-CoV-2 infection, as such the time since last infection is likely comparable between the two groups.

While we did not measure absolute VE against infection or severe disease compared to unvaccinated, it provides valuable insights on the relative performance of a protein-based vaccine platform compared to the mRNA platform and will help formulate public health strategies. Based on prior immunological studies and our findings, it is advisable to assess the overall benefits and risks of different vaccine platforms in the public’s best interest (6).

In conclusion, our findings provide needed insight on short-term effectiveness of different COVID-19 vaccine platforms. Furthermore, it underscores the importance for continued vigilance to ensure increased COVID-19 vaccination coverage perhaps through broader vaccine options.

## Data Availability

The authors do not have permission to share data.

## Acknowledgments

This study used the Korea Disease Control and Prevention Agency (KDCA) and National Health Insurance Service (NHIS) databases for policy and academic research (KDCA-NHIS-2023-1-494). The conclusions of this study are not related to this institution.

## Contributions

Conceptualization and study design: EG, SC, EB, YJC, CW, and MDR. Data acquisition and analysis: EG, SC, EB, and YJC. Data interpretation: EG, SC, EB, YJC, CW, JF, MV, and MDR. Drafting of the manuscript and critical review: EG, SC, EB, YJC, CW, JF, MV, and MDR.

